# Factors associated with discharge and return visits of emergency department patients with a sepsis best practice alert

**DOI:** 10.64898/2026.10.06.26364872

**Authors:** Benjamin Wyler, Chen Yang, Samuel E. Sondheim, Michael Lanigan, Nathan I. Shapiro, Madhu Mazumdar, Roland C. Merchant

## Abstract

**Background:** Best practice alerts (BPAs) used in emergency departments (EDs) to identify patients with possible sepsis lack specificity and some ED patients with sepsis BPAs are discharged. We evaluated factors associated with ED discharge following a sepsis BPA and with subsequent 7-day return admissions with sepsis or infection.

**Methods:** We conducted a retrospective cohort study using an electronic health record data query from five hospitals (2022-2024). Using logistic regression, we evaluated the association of participant demographics and clinical characteristics with ED discharge and factors at the initial ED visit associated with return admission.

**Results:** Among 62,068 ED visits with a sepsis BPA, one third resulted in ED discharge. Discharged participants were younger and more often female, Black, multiracial, and/or Hispanic; they more often had an initially normal pulse, respiratory rate, blood pressure, and lactic acid than admitted participants. Discharged participants more often had a fever on arrival (adjusted odds ratio (aOR) 6.03; p<0.001) and a white blood cell (WBC) count 11,000-14,999 (aOR 4.57; p<0.001) or 15,000-19,999/uL (aOR 2.01; p<0.001). One percent of discharged participants experienced a 7-day return admission with a diagnosis of sepsis or infection, which was associated with older age, White or Asian race, diastolic hypotension, WBC count ≤15,000/uL or <4,000/uL, and an index visit BPA response indicating sepsis or non-response.

**Conclusions:** Return admissions among discharged ED patients with a sepsis BPA are rare. Several factors are associated both with hospital admission at the index visit and return admission, indicating a need for additional risk-stratification tools.

## 1 INTRODUCTION

Sepsis is a life-threatening emergency resulting from a dysregulated host immune response to infection. Approximately 80% of patients with sepsis in the United States (US) receive initial care in the emergency department (ED). [1,2] Sepsis guidelines and a core measure (SEP-1) by the Centers for Medicare and Medicaid Services (CMS) have incentivized early treatment of patients with sepsis. [3,4] To meet these guidelines, EDs employ screening tools and best practice alerts (BPAs) linked to the electronic health record (EHR) and triggered by the presence of systemic inflammatory response syndrome (SIRS) to aid early identification and treatment of patients who might have sepsis. SIRS is estimated to occur in 17.6% of ED patients [5] and is a moderately sensitive indicator of infection, yet is has poor specificity for sepsis, because not all infections with SIRS have life-threatening immune dysregulation and many non-infectious conditions can cause SIRS. [6–9] Nonetheless, sepsis BPAs have been linked with improved mortality and shorter hospital length of stay (LOS). [1,10]

To comply with SEP-1, ED clinicians must decide within limited time frames whether to initiate sepsis treatment for patients who trigger a sepsis BPA. Unfortunately, they often have limited information upon which to base this decision and may rely on non-patient factors. [11] Not all ED patients with sepsis BPAs are treated empirically and admitted to the hospital, but research is limited on the frequency with which such patients are discharged from the ED and the factors that might influence physician decision making regarding ED admission or discharge. Additionally, there is limited data on the safety of ED discharge of patients with a sepsis BPA, including the frequency with which these patients return to the ED and are admitted for sepsis or an infection, or the factors that are associated with that outcome. However, some evidence suggests that even patients meeting the most up-to-date international consensus criteria for diagnosing sepsis, Sepsis-3, who are discharged from EDs generally have favorable outcomes, with 30-day mortality of less than 1%. [12]

Using an EHR data query, we sought to understand physician actions when faced with a sepsis BPA, including documented responses to the BPA that required classification of the patient as septic or not septic, patient diagnoses, and patient dispositions. We further sought to evaluate which patient factors (demographics, initial vital signs, laboratory test results) were associated with admission or discharge among ED patients with a sepsis BPA. We also assessed the frequency of 7-day return ED visits and admissions among these patients and sought to identify factors from the initial visit that could be associated with return admissions.

## 2 METHODS

### 2.1 Study Design, Population and Setting

This retrospective cohort study analyzed electronic health records (EHRs) data from adult patients who triggered a sepsis BPA between January 1, 2022 and December 31, 2024, across five XXX City hospitals (xxx) in the same healthcare system. The EDs at these hospitals had a combined total of 348,079 visits in 2024. The health system’s Internal Review Board (IRB) approved the study and granted a waiver of informed consent prior to commencement. The Strengthening the Reporting of Observational Studies in Epidemiology (STROBE) guidelines were followed to ensure appropriate reporting of this observational study (see Appendix Table A1). [13]

### 2.2 Study Participants

We included all ED patients who had a sepsis BPA within 4 hours of ED arrival across the 5 participating hospitals. The sepsis BPA could be triggered by: 1) SBP<90 on initial vital signs; 2) two or more SIRS criteria on initial vital signs (temperature >38C or <36C, pulse >90, respiratory rate >20, oxygen saturation <90%); 3) two or more SIRS criteria (temperature >38C or <36C, pulse >90, respiratory rate >20, oxygen saturation <90%, WBC count >12K or <4K) plus a lactic acid >2.0mg/dl or systolic BP<90 mmHg at any time after ED arrival. Patients who had a sepsis BPA more than 4 hours from ED arrival were excluded because their initial vital signs and laboratory test results might not directly affect initial clinician decision-making about sepsis treatments, or such patients likely were boarding in the ED after admission. We excluded patients with a recorded age ≥110 years, given that this age is assigned in the EHR for patients whose identity cannot be verified. For each unique patient medical record number (MRN), the index ED visit was defined as an encounter without a preceding ED visit within 7 days. Subsequent encounters by a patient with the same MRN occurring within 7 days of the index visit were defined as follow-up ED visits. Subsequent ED visits occurring more than 7 days after an index visit were considered new index ED visits.

### 2.3 Extracted Data for Analysis

We extracted EHR data on patient demographic characteristics (age, sex, race, insurance status, primary residence ZIP code), temporal variables (ED arrival date and time, ED and inpatient LOS), initial vital sign measurements (initial temperature, pulse, blood pressure, respiratory rate), first laboratory test result during the ED visit (WBC count, serum lactic acid, blood culture), sepsis BPA classification by the ED physician (sepsis, severe sepsis, septic shock, SIRS/other), ED disposition, primary ED diagnosis (see Appendix B for methods for categorization), any return ED visit within 7-days, and disposition and ED diagnosis during return visit.

### 2.4 Data Analysis

Statistical analyses were conducted using SAS® analytical software (SAS Institute, Inc., Cary, NC) and R (version 4.6.1). We calculated descriptive statistics for study sites and patient dispositions for all participants. We also generated descriptive statistics for participant demographic characteristics, BPA classifications, and ED diagnostic categories for all participants, stratified by ED disposition (admission (including operating room/procedure, transfer, or observation)), ED discharge, and all others (premature departure, expired in ED, transfer to labor and delivery, or EHR downtime chart)). Blood culture results were classified as pathogens, likely contaminants, or equivocal by the principal investigator following established standards. [14]

#### 2.4.1 Univariable and Multivariable Modeling

We developed univariable logistic regression models to evaluate the association between ED discharge (vs. hospital admission) and demographic characteristics, initial vital signs, and laboratory results. Predictors were categorized or dichotomized using usual clinical parameters and to ensure sufficient subgroup sizes for model stability. Variables achieving a significant threshold of p <0.20 in univariable analysis were advanced to multivariable logistic regression modeling.

#### 2.4.2 Frequency of 7-day Return ED Visits

To evaluate 7-day return ED visits, we restricted our analysis to participants whose residential ZIP codes matched the borough of the hospital site (Manhattan, Queens, or Brooklyn) where their index ED visit occurred. This approach was intended to eliminate potential bias from inclusion of patients who would be less likely to return to a XXX ED, including travelers and work commuters. We calculated the frequencies of 7-day return ED visit and return ED visit with hospital admission. To estimate the frequency of return ED visit admissions that were likely related to sepsis, we further stratified these cases by primary ED diagnoses that could be associated with sepsis (sepsis, infection, possible infection, and organ failure). Return admissions carrying a primary ED diagnostic category of sepsis or infection were considered indicative of a likely missed sepsis diagnosis during the index visit.

#### 2.4.3 Factors Associated with 7-day Return Admission with Sepsis or Infection ED Diagnosis

As an exploratory analysis, we sought to identify factors during the index ED visit that were associated with 7-day return admission with likely missed sepsis diagnosis. We generated univariable logistic regression models that related 7-day return hospitalization with primary ED diagnosis of sepsis or infection with patient-level factors (demographic and clinical characteristics), hospital site, clinician impressions (sepsis BPA classifications and ED diagnostic categories) and operational metrics (ED LOS and ED observation unit use), during the index visit. We also compared participants with 7-day return admissions for sepsis or infection to participants who were discharged at the index ED visit and either (1) had no 7-day return ED visit or (2) experienced a 7-day return ED visit that did not result in hospital admission. Multivariable analyses were not possible due to the large number of factors in comparison to the number of outcomes.

## 3 RESULTS

### 3.1 Participant sample, Demographics, Clinical Sites, and Sepsis BPA responses

Our EHR query identified 66,487 ED patient encounters with sepsis BPAs, with 49,350 unique patient MRNs. After excluding ED encounters with the same MRN that occurred within 7 days following an initial ED encounter (i.e., return visit, n=1,042); BPA times that were >4 hours from ED arrival (n=2,937), prior to ED arrival time (n=9), or missing (n=227); and patient age ≥110 years (n=218), there remained 62,068 index ED visits for final analysis. Participants with more than one index visit were common (n=7,562), 95.8% of whom had 2-5 visits during the study period.

The median participant age was 58 years (interquartile range 38-73). Nearly one in three participants were 70 years or older. There was a slight female predominance. There were similar proportions of participants who were White, Black, and Other race and over a quarter were Hispanic. Two-thirds of participants had Medicare or Medicaid health care insurance (Table 1).

**Table 1:** Participant Demographic Characteristics by ED Disposition shows the demographic characteristics of participants among 62,062 ED encounters with a sepsis BPA by number and proportion, both in aggregate and by ED disposition. Participant proportions by ED disposition are calculated by row.

|  | All patients | Discharged | Admitted* | Premature departure/other |
| --- | --- | --- | --- | --- |
| Age | n (%) | n (%) | n (%) | n (%) |
| 18-29 | 8633 (13.9) | 6137 (71.1) | 2190 (25.4) | 298 (3.5) |
| 30-39 | 7980 (12.9) | 4637 (58.1) | 2967 (37.2) | 365 (4.6) |
| 40-49 | 6827 (11.0) | 3155 (46.2) | 3374 (49.4) | 293 (4.3) |
| 50-59 | 8856 (14.3) | 3129 (35.2) | 5361 (60.5) | 352 (4.0) |
| 60-69 | 11138 (17.9) | 2701 (24.3) | 8025 (72.1) | 406 (3.6) |
| 70-79 | 9338 (15.0) | 1526 (16.3) | 7552 (80.9) | 255 (2.7) |
| 80-89 | 6642 (10.7) | 704 (10.6) | 5760 (86.7) | 174 (2.6) |
| 90-99 | 2523 (4.1) | 172 (6.8) | 2269 (89.9) | 81 (3.2) |
| 100-109 | 131 (0.2) | 7 (5.3) | 121 (92.4) | 3 (2.3) |
| Sex |  |  |  |  |
| Male | 30665 (49.4) | 9937 (32.4) | 19456 (63.4) | 1242 (4.1) |
| Female | 31397 (50.6) | 12229 (38.9) | 18160 (57.8) | 985 (3.1) |
| Unk/missing | 6 (0.0) | 3 (50.0) | 3 (50.0) | 0 (0.0) |
| Race |  |  |  |  |
| White | 16700 (26.9) | 4639 (27.8) | 11469 (68.7) | 596 (3.5) |
| Black | 19294 (31.1) | 6970 (36.1) | 11524 (59.7) | 782 (4.1) |
| Asian | 3272 (5.3) | 1142 (34.9) | 2054 (62.8) | 73 (2.2) |
| Other | 20379 (32.8) | 8481 (41.6) | 11196 (54.9) | 681 (3.3) |
| Multiracial | 175 (0.3) | 75 (42.9) | 90 (51.4) | 10 (5.7) |
| Unknown | 2236 (3.6) | 852 (38.1) | 1284 (57.4) | 95 (4.2) |
| Missing | 12 (0.0) | 10 (0.0) | 2 (0.0) | 0 (0.0) |
| Ethnicity |  |  |  |  |
| Hispanic | 17594 (28.3) | 7227 (41.1) | 9798 (55.7) | 580 (3.2) |
| non-Hispanic | 39176 (63.1) | 13082 (33.4) | 24597 (62.8) | 1535 (3.8) |
| Unknown/other | 5298 (8.5) | 1860 (35.1) | 3224 (60.9) | 213 (4.0) |
| Insurance |  |  |  |  |
| Medicare | 23648 (38.1) | 3921 (16.6) | 19028 (80.5) | 711 (3.0) |
| Medicaid | 18060 (29.1) | 7988 (44.2) | 9261 (51.3) | 839 (4.5) |
| Commercial | 10893 (17.6) | 5544 (50.9) | 5060 (46.5) | 290 (2.6) |
| Self-Pay | 7740 (12.5) | 3692 (47.7) | 3615 (46.7) | 442 (5.6) |
| Other | 1727 (2.8) | 1024 (59.3) | 655 (37.9) | 46 (2.6) |

\*Includes OR or procedure, observation, and transfer

The distribution of participant ED encounters varied greatly across hospital sites (Table 2). Physician responses to sepsis BPAs indicated that the sepsis was not suspected in more than half of ED visits, while sepsis was suspected in a third. Conversely, more than half of participants were admitted to the hospital and a third of participants were discharged.

**Table 2:** Frequency of Participants by Clinical Site, Sepsis Classification, and ED Disposition (n=62,068) shows the distribution of clinical site of encounter, the ED clinician classification in response to the sepsis BPA, the ED disposition by total numbers and proportions of participants among 62,062 ED encounters with a sepsis BPA.

| Clinical Site | n | % |
| --- | --- | --- |
| 1 | 6026 | 9.7 |
| 2 | 23219 | 37.4 |
| 3 | 15003 | 24.2 |
| 4 | 7816 | 12.6 |
| 5 | 10004 | 16.1 |
| BPA classification by ED clinician |  |  |
| No sepsis | 35178 | 56.7 |
| Any sepsis | 20735 | 33.4 |
| Sepsis | 17503 | 28.2 |
| Severe Sepsis | 2181 | 3.5 |
| Septic Shock | 1051 | 1.7 |
| Sepsis ruled out* | 49 | 0.2 |
| Not documented | 6155 | 9.9 |
| Disposition |  |  |
| Admission | 35305 | 56.9 |
| Operating room / procedure | 451 | 0.7 |
| Transfer | 1840 | 3.0 |
| Observation | 607 | 1.0 |
| Discharge | 21579 | 34.8 |
| Premature departure** | 1725 | 2.8 |
| Other or unknown | 41 | 0.1 |
| Missing | 53 | 0.1 |

\* Percentage denominator is all patients initially classified as sepsis
\*\* Includes left before treatment complete, left against medical advice, left without being seen, or eloped

### 3.2 Sepsis Classifications, ED Diagnoses, and ED LOS by Disposition

ED dispositions varied by sepsis BPA response and ED diagnosis (Table 3). Over half of participants classified as not having sepsis were discharged, while those classified as having sepsis were rarely discharged. Fewer than 1% of patients with an ED *diagnosis* of sepsis were discharged, while discharge rates among patients with infection diagnosis varied by infection source. Patients with non-infectious or possible infection ED diagnostic categories had lower rates of discharge than those with infection diagnosis. Discharged patients were more likely to have ED LOS <6 hours. Blood culture results among admitted and discharged participants are provided in the Appendix C.

**Table 3:** Sepsis BPA Classifications, ED Diagnostic Categories, and ED Length of Stay in Relation to ED Dispositions shows the distribution of sepsis classifications, ED diagnoses, and ED length of stay in total numbers and proportions of participants among 62,062 ED encounters with a sepsis BPA. Participant proportions by ED disposition are calculated by row.

|  | All patients |  | Discharged |  | Admitted* |  | Premature departure/other** |
| --- | --- | --- | --- | --- | --- | --- | --- |
| Sepsis classification | n | (%) | n | (%) | n | (%) | n (%) |
| No sepsis | 35178 | (56.7) | 18923 | (53.8) | 14593 | (41.5) | 1662 (4.7) |
| Any sepsis | 20735 | (33.4) | 2620 | (12.6) | 17643 | (85.1) | 472 (2.3) |
| Sepsis | 17503 | (28.2) | 2525 | (14.4) | 14600 | (83.4) | 378 (2.2) |
| Severe sepsis | 2181 | (3.5) | 82 | (3.8) | 2052 | (94.1) | 47 (2.1) |
| Septic shock | 1051 | (1.7) | 13 | (1.2) | 991 | (94.3) | 47 (4.5) |
| Sepsis ruled out | 49 | (0.2) | 11 | (22.4) | 38 | (77.6) | 0 (0.0) |
| Not documented | 6155 | (9.9) | 626 | (10.2) | 5383 | (87.5) | 143 (2.3) |
| ED diagnostic category |  |  |  |  |  |  |  |
| Sepsis | 3484 | (5.6) | 29 | (0.8) | 3400 | (97.6) | 54 (1.5) |
| Serious infection | 1576 | (2.5) | 131 | (8.3) | 1421 | (90.2) | 23 (1.5) |
| Infection | 18453 | (29.7) | 8664 | (47.0) | 9453 | (51.2) | 325 (1.9) |
| Pneumonia | 3705 | (6.0) | 724 | (19.5) | 2926 | (79.0) | 51 (1.5) |
| GU infection | 2323 | (3.7) | 781 | (33.6) | 1515 | (65.2) | 27 (1.2) |
| SSTI | 1412 | (2.3) | 378 | (26.8) | 978 | (69.3) | 55 (4.0) |
| GI infection | 911 | (1.5) | 462 | (50.7) | 434 | (47.6) | 13 (1.4) |
| ENT infection | 1191 | (1.9) | 1027 | (86.2) | 144 | (12.1) | 19 (1.6) |
| Febrile Syndrome | 70 | (0.1) | 38 | (54.3) | 28 | (40.0) | 4 (5.7) |
| Other/unspecified | 1616 | (2.6) | 708 | (43.8) | 837 | (51.8) | 70 (4.3) |
| Viral infection | 5649 | (9.1) | 4415 | (78.2) | 1170 | (20.7) | 63 (1.1) |
| Possible infection | 6814 | (11.0) | 2161 | (31.7) | 4364 | (64.0) | 284 (4.3) |
| Non-infectious | 29216 | (47.1) | 11138 | (38.1) | 16695 | (57.1) | 1352 (4.7) |
| Organ failure | 3676 | (5.9) | 133 | (3.6) | 3461 | (94.2) | 80 (2.3) |
| ED length of stay |  |  |  |  |  |  |  |
| <6 hours | 29216 | (47.1) | 11138 | (38.1) | 16695 | (57.1) | 1081 (6.2) |
| ≥6 hours | 3676 | (5.9) | 133 | (3.6) | 3461 | (94.2) | 1199 (2.7) |

\* Includes OR, procedure, observation, and transfer
\*\* Includes premature departure, sent to L&D, expired in ED, downtime chart

### 3.3 Demographic and Clinical Factors Associated with ED Discharge

From the results of the multivariable analysis (Table 4), factors from the index ED visit associated with greater odds of ED discharge were younger age, non-White/non-Asian race, Hispanic ethnicity, female sex, initial temperature 38.0-39.9C, WBC count 11,000-19,999/uL, and missing data for WBC count and/or lactic acid (likely due to lack of testing). Factors associated with lower odds of ED discharge were initial hypotension, initial pulse ≥90, initial tachypnea, WBC count <4,000/mL or ≥20,000/mL, and lactic acid >2.0mmol/dL.

**Table 4:**
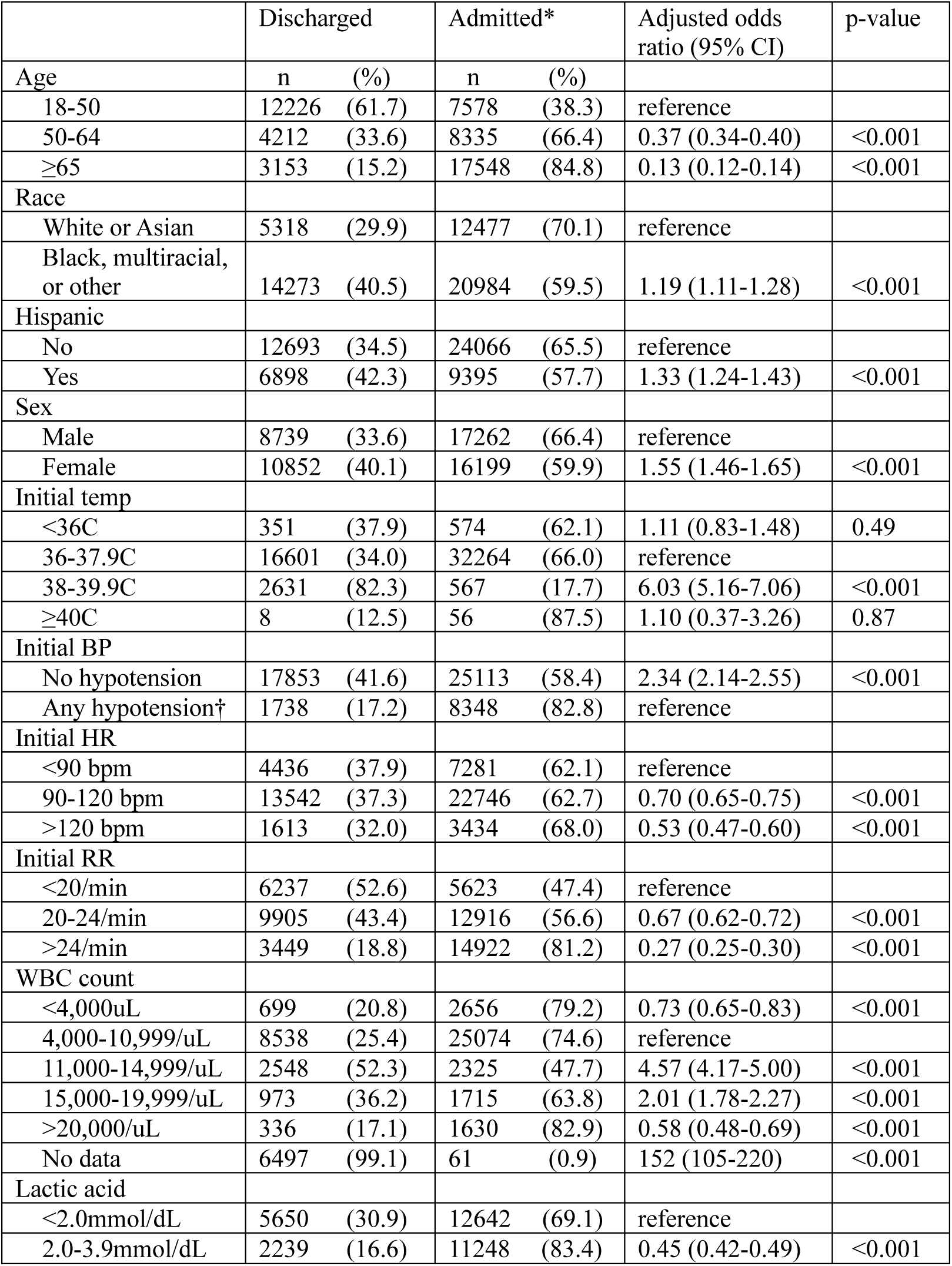
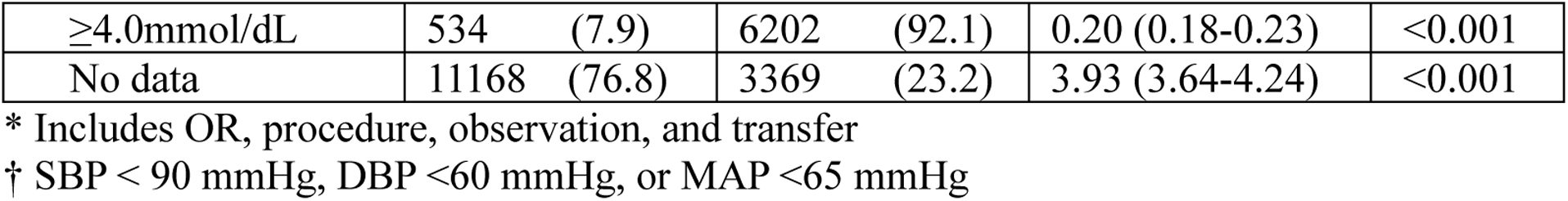
Multivariable Analysis of Factors Associated with ED Discharge of Patients who Triggered a Sepsis BPA shows the numbers and proportions of participants among 62,062 ED encounters with a sepsis BPA who were admitted or discharged from the ED, based upon demographic and clinical data found to be associated with ED disposition in univariable analysis. It also shows the adjusted odds ratios for ED discharge, 95% confidence intervals, and p-values for each covariate found in multivariable logistic regression analysis.

|  | Discharged |  | Admitted* |  | Adjusted odds ratio (95% CI) | p-value |
| --- | --- | --- | --- | --- | --- | --- |
| Age | n | (%) | n | (%) |  |  |
| 18-50 | 12226 | (61.7) | 7578 | (38.3) | reference |  |
| 50-64 | 4212 | (33.6) | 8335 | (66.4) | 0.37 (0.34-0.40) | <0.001 |
| $\geq 65$ | 3153 | (15.2) | 17548 | (84.8) | 0.13 (0.12-0.14) | <0.001 |
| Race |  |  |  |  |  |  |
| White or Asian | 5318 | (29.9) | 12477 | (70.1) | reference |  |
| Black, multiracial, or other | 14273 | (40.5) | 20984 | (59.5) | 1.19 (1.11-1.28) | <0.001 |
| Hispanic |  |  |  |  |  |  |
| No | 12693 | (34.5) | 24066 | (65.5) | reference |  |
| Yes | 6898 | (42.3) | 9395 | (57.7) | 1.33 (1.24-1.43) | <0.001 |
| Sex |  |  |  |  |  |  |
| Male | 8739 | (33.6) | 17262 | (66.4) | reference |  |
| Female | 10852 | (40.1) | 16199 | (59.9) | 1.55 (1.46-1.65) | <0.001 |
| Initial temp |  |  |  |  |  |  |
| $< 36^{\circ}\text{C}$ | 351 | (37.9) | 574 | (62.1) | 1.11 (0.83-1.48) | 0.49 |
| $36-37.9^{\circ}\text{C}$ | 16601 | (34.0) | 32264 | (66.0) | reference | |
| $38-39.9^{\circ}\text{C}$ | 2631 | (82.3) | 567 | (17.7) | 6.03 (5.16-7.06) | <0.001 |
| $\geq 40^{\circ}\text{C}$ | 8 | (12.5) | 56 | (87.5) | 1.10 (0.37-3.26) | 0.87 |
| Initial BP |  |  |  |  |  |  |
| No hypotension | 17853 | (41.6) | 25113 | (58.4) | 2.34 (2.14-2.55) | <0.001 |
| Any hypotension† | 1738 | (17.2) | 8348 | (82.8) | reference |  |
| Initial HR |  |  |  |  |  |  |
| $< 90$ bpm | 4436 | (37.9) | 7281 | (62.1) | reference | |
| 90-120 bpm | 13542 | (37.3) | 22746 | (62.7) | 0.70 (0.65-0.75) | <0.001 |
| $> 120$ bpm | 1613 | (32.0) | 3434 | (68.0) | 0.53 (0.47-0.60) | <0.001 |
| Initial RR |  |  |  |  |  |  |
| $< 20/\text{min}$ | 6237 | (52.6) | 5623 | (47.4) | reference | |
| 20-24/min | 9905 | (43.4) | 12916 | (56.6) | 0.67 (0.62-0.72) | <0.001 |
| $> 24/\text{min}$ | 3449 | (18.8) | 14922 | (81.2) | 0.27 (0.25-0.30) | <0.001 |
| WBC count |  |  |  |  |  |  |
| $< 4,000/\text{uL}$ | 699 | (20.8) | 2656 | (79.2) | 0.73 (0.65-0.83) | <0.001 |
| 4,000-10,999/uL | 8538 | (25.4) | 25074 | (74.6) | reference |  |
| 11,000-14,999/uL | 2548 | (52.3) | 2325 | (47.7) | 4.57 (4.17-5.00) | <0.001 |
| 15,000-19,999/uL | 973 | (36.2) | 1715 | (63.8) | 2.01 (1.78-2.27) | <0.001 |
| $> 20,000/\text{uL}$ | 336 | (17.1) | 1630 | (82.9) | 0.58 (0.48-0.69) | <0.001 |
| No data | 6497 | (99.1) | 61 | (0.9) | 152 (105-220) | <0.001 |
| Lactic acid |  |  |  |  |  |  |
| $< 2.0\text{mmol/dL}$ | 5650 | (30.9) | 12642 | (69.1) | reference | |
| 2.0-3.9mmol/dL | 2239 | (16.6) | 11248 | (83.4) | 0.45 (0.42-0.49) | <0.001 |

|  |  |  |  |  |
| --- | --- | --- | --- | --- |
| ≥4.0mmol/dL | 534 (7.9) | 6202 (92.1) | 0.20 (0.18-0.23) | <0.001 |
| No data | 11168 (76.8) | 3369 (23.2) | 3.93 (3.64-4.24) | <0.001 |

\* Includes OR, procedure, observation, and transfer
† SBP < 90 mmHg, DBP <60 mmHg, or MAP <65 mmHg

### 3.4 7-day Return Visits, Admissions, and Associated Diagnoses

Of the 21,579 participants who were discharged at the index ED visit, 5,830 (27.0%) resided in a different borough than the ED they visited. Of the remaining 15,749 participants who were discharged from the index ED visit and resided in the same borough as the ED they visited, 1,750 (11.1%) returned to an ED in the health system within 7 days of their index ED visit, and 568 (3.6%) were admitted at a return ED visit: 0.1% with an ED diagnosis of sepsis and 0.9% with infection, resulting in an estimated frequency of a possible missed sepsis diagnosis of 1.0%.

### 3.5 Factors Associated with 7-day Return Admission for Sepsis or Infection

Among 15,749 participants discharged at the index ED visit who resided in the same borough as the ED they visited, factors from the index ED visit associated with greater odds of a possible missed sepsis diagnosis, as compared to either no return ED visit within 7-days or no admission at a return ED visit within 7 days are shown in Table 5. Missing data for WBC count and lactic acid, likely reflecting that testing was not performed, was associated with lower odds of possible missed sepsis diagnosis.

**Table 5:** Univariable Analysis of Factors Associated with 7-day Return ED Visit and Admission with ED Diagnosis of Sepsis or Infection compares the characteristics of 157 participants who were discharged at an initial ED visit with a sepsis BPA and had a return visit within 7 days with a hospital admission and diagnosis of sepsis or infection to 15181 participants who did not have that outcome. Univariable odds ratios, confidence intervals, and p-values are provided for the correlation of each participant characteristic with these outcomes.

|  | Possible missed sepsis† |  | All others |  | Odds ratio (95% CI) | p-value |
| --- | --- | --- | --- | --- | --- | --- |
| Age | n | (%) | n | (%) |  |  |
| 18-50 | 60 | (0.6) | 9521 | (99.4) | reference |  |
| 50-64 | 45 | (1.3) | 3290 | (98.7) | 2.17 (1.47-3.20) | <0.001 |
| ≥65 | 52 | (1.8) | 2781 | (98.2) | 2.97 (2.04-4.31) | <0.001 |
| Race |  |  |  |  |  |  |
| White or Asian | 58 | (1.5) | 3881 | (98.5) | reference |  |
| Black, multiracial or other | 96 | (0.9) | 11109 | (99.1) | 0.58 (0.42-0.80) | 0.001 |
| Hispanic |  |  |  |  |  |  |
| No | 94 | (1.0) | 9092 | (99.0) | reference |  |
| Yes | 57 | (1.1) | 5180 | (98.9) | 1.06 (0.76-1.48) | 0.71 |
| Sex |  |  |  |  |  |  |
| Male | 70 | (1.0) | 6915 | (99.0) | reference |  |
| Female | 87 | (1.0) | 8676 | (99.0) | 0.99 (0.72-1.36) | 0.95 |
| Initial temp |  |  |  |  |  |  |
| <36C | 1 | (0.4) | 283 | (99.6) | 0.34 (0.05-2.45) | 0.29 |
| 36-37.9C | 135 | (1.0) | 13046 | (99.0) | reference |  |
| 38-39.9C | 21 | (1.0) | 2077 | (99.0) | 0.98 (0.62-1.55) | 0.92 |
| ≥40C | 0 | (0.0) | 7 | (100.0) | n/a | >0.99 |
| Initial BP* |  |  |  |  |  |  |
| SBP<90mmHg | 6 | (1.4) | 428 | (98.6) | 1.40 (0.61-3.17) | 0.43 |
| DBP<60mmHg | 27 | (2.0) | 1331 | (98.0) | 2.20 (1.45-3.35) | <0.001 |
| MAP<65mmHg | 7 | (1.9) | 355 | (98.1) | 1.99 (0.92-4.27) | 0.079 |
| Initial HR |  |  |  |  |  |  |
| <90 bpm | 45 | (1.2) | 3572 | (98.8) | reference |  |
| 90-120 bpm | 95 | (0.9) | 10654 | (99.1) | 0.71 (0.50-1.01) | 0.058 |
| >120 bpm | 17 | (1.3) | 1293 | (98.7) | 1.04 (0.60-1.83) | 0.88 |
| Initial RR |  |  |  |  |  |  |
| <20/min | 57 | (1.1) | 5213 | (98.9) | reference |  |
| 20-24/min | 74 | (1.0) | 7604 | (99.0) | 0.89 (0.63-1.26) | 0.51 |
| >24/min | 26 | (1.0) | 2648 | (99.0) | 0.90 (0.56-1.43) | 0.65 |
| WBC count |  |  |  |  |  |  |
| <4,000/uL | 11 | (2.1) | 507 | (97.9) | 2.26 (1.19-4.32) | 0.013 |
| 4,000-10,999/uL | 65 | (0.9) | 6781 | (99.1) | reference |  |
| 11,000-14,999/uL | 25 | (1.2) | 2078 | (98.8) | 1.26 (0.79-2.00) | 0.34 |
| 15,000-19,999/uL | 21 | (2.7) | 760 | (97.3) | 2.88 (1.75-4.74) | <0.001 |
| >20,000/uL | 9 | (3.2) | 270 | (96.8) | 3.48 (1.71-7.06) | <0.001 |
| No data | 26 | (0.5) | 5196 | (99.5) | 0.52 (0.33-0.82) | 0.005 |
| Lactic acid |  |  |  |  |  |  |
| <2.0mmol/dL | 69 | (1.5) | 4528 | (98.5) | reference |  |
| 2.0-3.9mmol/dL | 31 | (1.7) | 1801 | (98.3) | 1.13 (0.74-1.73) | 0.58 |
| ≥4.0mmol/dL | 2 | (0.4) | 450 | (99.6) | 0.29 (0.07-1.19) | 0.087 |
| No data | 45 | (0.6) | 3572 | (99.4) | 0.41 (0.29-0.58) | <0.001 |
| Index visit sepsis BPA classification |  |  |  |  |  |  |
| Sepsis | 62 | (3.2) | 1903 | (96.8) | 5.01 (3.60-6.98) | <0.001 |
| Not sepsis | 86 | (0.6) | 13236 | (99.4) | reference |  |
| No response | 9 | (1.9) | 453 | (98.1) | 3.06 (1.53-6.11) | 0.002 |
| Index visit ED diagnosis |  |  |  |  |  |  |
| Sepsis | 2 | (10.5) | 17 | (89.5) | 15.8 (3.56-69.8) | <0.001 |
| Serious infection | 6 | (6.0) | 94 | (94.0) | 8.55 (3.60-20.3) | <0.001 |
| Infection | 80 | (1.3) | 6186 | (98.7) | 1.73 (1.23-2.44) | 0.002 |
| Pneumonia | 12 | (2.3) | 505 | (97.7) | 3.18 (1.70-5.97) | <0.001 |
| GU infection | 18 | (3.2) | 537 | (96.8) | 4.49 (2.62-7.68) | <0.001 |
| SSTI | 3 | (1.1) | 265 | (98.9) | 1.52 (0.47-4.87) | 0.48 |
| GI infection | 3 | (0.9) | 326 | (99.1) | 1.23 (0.38-3.96) | 0.73 |
| ENT infection | 6 | (0.8) | 756 | (99.2) | 1.06 (0.46-2.47) | 0.89 |
| Febrile syndrome | 1 | (3.8) | 25 | (96.2) | 5.36 (0.71-40.2) | 0.10 |
| Other infection | 20 | (4.0) | 481 | (96.0) | 5.57 (3.32-9.34) | <0.001 |
| Viral infection | 17 | (0.5) | 3291 | (99.5) | 0.69 (0.40-1.19) | 0.18 |
| Possible infection | 10 | (0.6) | 1554 | (99.4) | 0.86 (0.44-1.69) | 0.66 |
| Organ failure | 2 | (2.4) | 83 | (97.6) | 3.23 (0.77-13.4) | 0.11 |
| Non-infectious | 57 | (0.7) | 7632 | (99.3) | reference |  |
| ED LOS |  |  |  |  |  |  |
| <6 hours | 56 | (0.7) | 8410 | (99.3) | 0.47 (0.34-0.66) | <0.001 |
| ≥6 hours | 101 | (1.4) | 7182 | (98.6) | reference |  |
| Hospital site |  |  |  |  |  |  |
| 1 | 37 | (0.7) | 5135 | (99.3) | reference |  |
| 2 | 27 | (1.1) | 2530 | (98.9) | 1.48 (0.90-2.44) | 0.12 |
| 3 | 42 | (0.9) | 4509 | (99.1) | 1.29 (0.83-2.01) | 0.26 |
| 4 | 38 | (1.6) | 2311 | (98.4) | 2.28 (1.45-3.60) | <0.001 |
| 5 | 13 | (1.2) | 1107 | (98.8) | 1.63 (0.86-3.08) | 0.13 |
| ED observation before discharge? |  |  |  |  |  |  |
| Yes | 3 | (0.8) | 368 | (99.2) | 0.81 (0.26-2.54) | 0.71 |
| No | 154 | (1.0) | 15224 | (99.0) | reference |  |

† Return visit within 7 days with ED diagnosis of sepsis or infection. Restricted to index visits that occurred in the same ZIP code as the patient's recorded residence
†† Comparator includes participants who were discharged at the index ED visit and did not have a 7-day return ED visit, or had a 7-day return ED visit that did not result in admission, transfer to another hospital, transfer to OR or procedural area, observation, or expired in the ED
\* Each category is binary, comparator is lack of condition

## 4 DISCUSSION

ED clinicians at the five hospitals in our study frequently deemed that patients with sepsis BPAs did not have sepsis (56.5%), and discharged over one third (34.7%), including 12.6% of those initially considered to have sepsis. Discharged patients tended to be younger, female, Black, multiracial, or other race, and Hispanic; had a shorter ED LOS, an ED diagnosis of infection (without sepsis), and clinical signs associated with infection (temperature 38.0-39.9C and WBC count 11,000-19,999/uL). Patients with a non-infectious ED diagnosis were discharged less often than those with an infection diagnosis, which suggests that many patients with sepsis BPAs may actually have either a mild infection or severe non-infectious condition. A sepsis BPA may therefore contribute to the risk of anchoring bias on sepsis, as well as overtreatment in non-infected patients. These risks were highlighted in a study by Shappell et. al. that found that ED patients who were treated for sepsis but found on post-hoc analysis to be unlikely to have bacterial infection had higher inpatient mortality rates compared with patients with high likelihood of bacterial infection (9.0% vs. 4.9%) and had significant rates of antibiotic harm (14.8%). [15]

Return visits to the ED within 7 days of discharge after a sepsis BPA were common at these five hospitals, occurring in 11.1% of discharged ED patients. This proportion of return visits is lower than the 25% rate observed by Moskowitz et. al. within 30 days among 179 ED patients with SIRS2+ who received intravenous (IV) antibiotics before being discharged, [16] which may be due to the shorter time window we studied and/or the lack of restriction to patients who received IV antibiotics. We deemed it likely that a discharged patient with sepsis would return to the ED within 7 days; however, a discharged patient might return for a variety of reasons unrelated to a missed sepsis diagnosis. We therefore focused on patients with features at the return visit that would likely occur in patients with a missed sepsis diagnosis—admission to the hospital and diagnosis of either sepsis or infection—which identified 1.0% of initially discharged participants as having a possible missed sepsis diagnosis, which rises to 2.1% with inclusion of participants diagnosed with possible infection or organ failure at the return visit. This admission rate is similar to the 1.11% rate of 7-day return sepsis admission reported in Chen et. al.’s study of patients discharged from the ED after pneumonia, UTI, and/or cellulitis. [17] Further investigation is needed to better measure infection-related outcomes after ED discharge among ED patients with a sepsis BPA.

We found that Asian, White, and older patients more often had a possible missed sepsis diagnosis. Associations between age and sepsis risk are well established, with increased risk due to higher comorbidity burden, immune senescence, and other factors. [18,19] On the other hand, previous literature has suggested that Black patients may be underdiagnosed and undertreated for sepsis, [20,21] which makes the current finding surprising.

We also found that initial diastolic hypotension and WBC counts <4,000/uL or ≥15,000/uL were associated with a possible missed sepsis diagnosis, which suggests that caution is warranted when considering discharge in patients with these clinical features. Previous studies of return sepsis admissions after ED visit have identified male sex, comorbidities (hypertension, chronic kidney disease, cardiovascular disease, cancer), higher temperature and heart rate, altered mental status, and electrolyte disturbances. [17,22] Further investigation using larger datasets that include additional clinical data might elucidate which factors are useful to use to reduce the risk of missed sepsis diagnosis at an ED visit with a sepsis BPA.

We found that ED patients are more likely to have a missed sepsis diagnosis if they are classified as having sepsis in response to the BPA, which usually occurs early in the clinical course. BPA non-response also is associated with likely missed sepsis diagnosis. These findings suggest that there may be risk in abandoning an initial clinical impression of sepsis, or neglecting to respond to the BPA, whether due to uncertainty, inadequate evaluation for sepsis, or another reason. Closer examination is warranted of the rare cases when ED patients are diagnosed with sepsis and discharged, as these cases were associated with return admission for sepsis or infection. Selective use of recently approved diagnostic tests with high negative predictive value for sepsis or sepsis-related outcomes, e.g. Intellisep (Cytovale, San Francisco, CA) and Triverity (Inflammatix, Sunnyvale, CA) may offer promise for further screening of patients with features associated with higher risk of missed sepsis diagnosis prior to discharge.

### 4.1 Limitations

Our study has several limitations. First, our assessment of possible missed sepsis diagnoses based on return ED visits with admission and ED diagnosis of sepsis or infection could lack accuracy as compared to Sepsis-3 criteria, which could have overestimated or underestimated the true proportion. It is also possible that the reason for admission on a return ED visit could have been unrelated to the severity of a diagnosed infection.

Second, our methodology could have missed return visits to EDs that occurred outside of the health system and/or out-of-hospital deaths, leading to underestimation of the frequency of missed sepsis diagnosis. Third, our ED diagnosis categories were assigned by the principal investigator using a schema that has not been previously validated and should be interpreted as exploratory. Fourth, we did not account for patient comorbidities, presenting symptoms, or antibiotic treatment, which could have been important factors in patient return visits. Fifth, as noted previously, because the proportion of discharged patients experiencing 7-day return admission for sepsis or infection was low, we could not perform multivariable analysis. Finally, although our study was drawn from a large, diverse patient population at multiple clinical sites, it may not be generalizable to other clinical settings. Differences in patient populations, practice patterns, and implementation and use of sepsis BPAs could lead to different results.

## 5 CONCLUSION

ED patients who trigger a sepsis BPA and are discharged are unlikely to have a missed sepsis diagnosis, with only 1.0% returning within 7-days and being admitted with a diagnosis of sepsis or infection. Many of the same factors that are associated with hospital admission during the initial presentation are also associated with return admissions among discharged patients, although moderate leukocytosis is associated with higher odds of both discharge and return admission. While current practices appear to carry a low risk of missed sepsis diagnosis, additional risk-stratification strategies are needed to help improve safe ED discharges following a sepsis BPA.

## Data Availability

All data produced in the present study are available upon reasonable request to the authors.

## Acknowledgements

Cindy Mei, MS assisted with performing the electronic health record data query.

## Author contributions

Study concept and design (BW, SES, ML, NIS, RCM); Acquisition of data (BW, CY); Analysis and interpretation of the data (BW, CY, MM, RCM); Drafting of the manuscript (BW); Critical revision of the manuscript for important intellectual content (SES, ML, NIS, MM, RCM); Statistical expertise (CY, MM); Acquisition of funding (N/A)

## REFERENCES

1. Yealy DM, Mohr NM, Shapiro NI, Venkatesh A, Jones AE, Self WH. Early Care of Adults With Suspected Sepsis in the Emergency Department and Out-of-Hospital Environment: A Consensus-Based Task Force Report. Ann Emerg Med. Jul 2021;78(1):1–19. doi:10.1016/j.annemergmed.2021.02.006

2. Wang HE, Jones AR, Donnelly JP. Revised National Estimates of Emergency Department Visits for Sepsis in the United States. Crit Care Med. Sep 2017;45(9):1443–1449. doi:10.1097/ccm.0000000000002538

3. Evans L, Rhodes A, Alhazzani W, et al. Surviving Sepsis Campaign: International Guidelines for Management of Sepsis and Septic Shock 2021. Critical Care Medicine. 2021;49(11):e1063–e1143. doi:10.1097/ccm.0000000000005337

4. Sloan SNB, Rodriguez N, Seward T, et al. Compliance with SEP-1 guidelines is associated with improved outcomes for septic shock but not for severe sepsis. J Intensive Med. Jul 2022;2(3):167–172. doi:10.1016/j.jointm.2022.03.003

5. Horeczko T, Green JP, Panacek EA. Epidemiology of the Systemic Inflammatory Response Syndrome (SIRS) in the emergency department. West J Emerg Med. May 2014;15(3):329–36. doi:10.5811/westjem.2013.9.18064

6. Gando S, Shiraishi A, Abe T, et al. The SIRS criteria have better performance for predicting infection than qSOFA scores in the emergency department. Sci Rep. May 15 2020;10(1):8095. doi:10.1038/s41598-020-64314-8

7. Oi Y, Ogawa F, Honzawa H, Abe T, Imaki S, Takeuchi I. A Screening Tool to Predict Sepsis in Patients With Suspected Infection in the Emergency Department. Cureus. Feb 2025;17(2):e78728. doi:10.7759/cureus.78728

8. van der Woude SW, van Doormaal FF, Hutten BA, F JN, Holleman F. Classifying sepsis patients in the emergency department using SIRS, qSOFA or MEWS. Neth J Med. May 2018;76(4):158–166.

9. Fernando SM, Tran A, Taljaard M, Cheng W, Rochwerg B, Seely AJE, Perry JJ. Prognostic Accuracy of the Quick Sequential Organ Failure Assessment for Mortality in Patients With Suspected Infection: A Systematic Review and Meta-analysis. Ann Intern Med. Feb 20 2018;168(4):266–275. doi:10.7326/m17-2820

10. Kim HJ, Ko RE, Lim SY, Park S, Suh GY, Lee YJ. Sepsis Alert Systems, Mortality, and Adherence in Emergency Departments: A Systematic Review and Meta-Analysis. JAMA Netw Open. Jul 1 2024;7(7):e2422823. doi:10.1001/jamanetworkopen.2024.22823

11. Souganidis ES, Patel B, Sampayo EM. Physician-Specific Utilization of an Electronic Best Practice Alert for Pediatric Sepsis in the Emergency Department. Pediatr Emerg Care. Aug 1 2022;38(8):e1417–e1422. doi:10.1097/pec.0000000000002778

12. Peltan ID, McLean SR, Murnin E, et al. Prevalence, Characteristics, and Outcomes of Emergency Department Discharge Among Patients With Sepsis. JAMA Netw Open. Feb 1 2022;5(2):e2147882. doi:10.1001/jamanetworkopen.2021.47882

13. von Elm E, Altman DG, Egger M, Pocock SJ, Gøtzsche PC, Vandenbroucke JP. The Strengthening the Reporting of Observational Studies in Epidemiology (STROBE) statement: guidelines for reporting observational studies. J Clin Epidemiol. Apr 2008;61(4):344–9. doi:10.1016/j.jclinepi.2007.11.008

14. Doern GV, Carroll KC, Diekema DJ, Garey KW, Rupp ME, Weinstein MP, Sexton DJ. Practical Guidance for Clinical Microbiology Laboratories: A Comprehensive Update on the Problem of Blood Culture Contamination and a Discussion of Methods for Addressing the Problem. Clin Microbiol Rev. Dec 18 2019;33(1)doi:10.1128/cmr.00009-19

15. Shappell CN, Yu T, Klompas M, et al. Frequency of Antibiotic Overtreatment and Associated Harms in Patients Presenting With Suspected Sepsis to the Emergency Department: A Retrospective Cohort Study. Clin Infect Dis. Jul 18 2025;80(6):1197–1207. doi:10.1093/cid/ciaf118

16. Moscowitz AE, Bae EY, Hoz RML, Cutrell JB, Monogue M. 62. Factors Associated with 30-Day ED Readmission Following Initial ED Discharge for Suspected Sepsis. Open Forum Infect Dis. Oct 2020;7(Suppl 1):S51. doi:10.1093/ofid/ofaa439.107

17. Chen AY, Allison M, Puskarich M, et al. Risk factors associated with return sepsis admission following emergency department discharge with infection. Am J Emerg Med. Nov 2025;97:207–215. doi:10.1016/j.ajem.2025.07.059

18. Qin C, Lin S, Pang L, et al. Risk factors for sepsis: a systematic review and meta-analysis. Review. Frontiers in Public Health. 2026-June-26 2026;Volume 14 - 2026doi:10.3389/fpubh.2026.1790141

19. Alhamyani AH, Alamri MS, Aljuaid NW, et al. Sepsis in Aging Populations: A Review of Risk Factors, Diagnosis, and Management. Cureus. Dec 2024;16(12):e74973. doi:10.7759/cureus.74973

20. Liu H, McCroskery S, Rajasekaran V, Linker AS, Poeran J, Truong TT. Racial Disparities in Documented Chief Complaints and Diagnoses in Sepsis Patients. J Intensive Care Med. Jul 2023;38(7):630–634. doi:10.1177/08850666231155397

21. Lizza BD, Betthauser KD, Juang PH, Hampton NB, Lyons PG, Kollef MH, Micek ST. Racial Disparities in Readmissions Following Initial Hospitalization for Sepsis. Crit Care Med. Mar 1 2021;49(3):e258–e268. doi:10.1097/ccm.0000000000004809

22. Horberg MA, Nassery N, Rubenstein KB, et al. Rate of sepsis hospitalizations after misdiagnosis in adult emergency department patients: a look-forward analysis with administrative claims data using Symptom-Disease Pair Analysis of Diagnostic Error (SPADE) methodology in an integrated health system. Diagnosis (Berl). Nov 25 2021;8(4):479–488. doi:10.1515/dx-2020-0145

23. Mellhammar L, Elén S, Ehrhard S, et al. New, Useful Criteria for Assessing the Evidence of Infection in Sepsis Research. Crit Care Explor. May 2022;4(5):e0697. doi:10.1097/cce.0000000000000697

