## Appendices for "Factors associated with discharge and return visits of emergency department patients with a sepsis best practice alert"

### Appendix A: STROBE Statement

Table A1: STROBE Statement—Checklist of Items that Should be Included in Reports of ***Cohort Studies***

| | Item No | Recommendation | Page No |
|---|---|---|---|
| **Title and abstract** | 1 | (*a*) Indicate the study's design with a commonly used term in the title or the abstract<br>(*b*) Provide in the abstract an informative and balanced summary of what was done and what was found | 1-4 |
| **Introduction** | | | |
| Background/rationale | 2 | Explain the scientific background and rationale for the investigation being reported | 5 |
| Objectives | 3 | State specific objectives, including any prespecified hypotheses | 5 |
| **Methods** | | | |
| Study design | 4 | Present key elements of study design early in the paper | 6 |
| Setting | 5 | Describe the setting, locations, and relevant dates, including periods of recruitment, exposure, follow-up, and data collection | 6 |
| Participants | 6 | (*a*) Give the eligibility criteria, and the sources and methods of selection of participants. Describe methods of follow-up<br>(*b*) For matched studies, give matching criteria and number of exposed and unexposed | 7 |
| Variables | 7 | Clearly define all outcomes, exposures, predictors, potential confounders, and effect modifiers. Give diagnostic criteria, if applicable | 8-9 |
| Data sources/ measurement | 8 | For each variable of interest, give sources of data and details of methods of assessment (measurement). Describe comparability of assessment methods if there is more than one group | 7-8 |
| Bias | 9 | Describe any efforts to address potential sources of bias | n/a |
| Study size | 10 | Explain how the study size was arrived at | 7 |
| Quantitative variables | 11 | Explain how quantitative variables were handled in the analyses. If applicable, describe which groupings were chosen and why | 8 |
| Statistical methods | 12 | (*a*) Describe all statistical methods, including those used to control for confounding<br>(*b*) Describe any methods used to examine subgroups and interactions<br>(*c*) Explain how missing data were addressed<br>(*d*) If applicable, explain how loss to follow-up was addressed<br>(*e*) Describe any sensitivity analyses | 9-10 |
| **Results** | | | |
| Participants | 13 | (a) Report numbers of individuals at each stage of study—eg numbers potentially eligible, examined for eligibility, confirmed eligible, included in the study, completing follow-up, and analysed<br>(b) Give reasons for non-participation at each stage<br>(c) Consider use of a flow diagram | 11 |
| Descriptive data | 14 | (a) Give characteristics of study participants (eg demographic, clinical, social) and information on exposures and potential confounders<br>(b) Indicate number of participants with missing data for each variable of interest | 11-12 |

| | | | |
|---|---|---|---|
| | | (c) Summarise follow-up time (eg, average and total amount) | |
| Outcome data | 15 | Report numbers of outcome events or summary measures over time | 12-14 |
| Main results | 16 | (*a*) Give unadjusted estimates and, if applicable, confounder-adjusted estimates and their precision (eg, 95% confidence interval). Make clear which confounders were adjusted for and why they were included<br>(*b*) Report category boundaries when continuous variables were categorized<br>(*c*) If relevant, consider translating estimates of relative risk into absolute risk for a meaningful time period | 12-14 |
| Other analyses | 17 | Report other analyses done—eg analyses of subgroups and interactions, and sensitivity analyses | n/a |
| **Discussion** | | | |
| Key results | 18 | Summarise key results with reference to study objectives | 14-17 |
| Limitations | 19 | Discuss limitations of the study, taking into account sources of potential bias or imprecision. Discuss both direction and magnitude of any potential bias | 17-18 |
| Interpretation | 20 | Give a cautious overall interpretation of results considering objectives, limitations, multiplicity of analyses, results from similar studies, and other relevant evidence | 18 |
| Generalisability | 21 | Discuss the generalisability (external validity) of the study results | 18 |
| **Other information** | | | |
| Funding | 22 | Give the source of funding and the role of the funders for the present study and, if applicable, for the original study on which the present article is based | n/a |

Table A1 shows the page numbers where different items that are considered standard for reporting in a cohort study, according to STROBE guidelines, are located in the manuscript.

### Appendix B: Classification of ED Diagnoses

To facilitate the analysis of ED primary diagnosis, the study's principal investigator prepared a classification system to group primary ED diagnoses, using as a model the schema described by Linden and Mellhammar. Their schema uses 13 foci of infection and classifies the likelihood of infection on a scale of 1-4. [23] Our classification system used seven primary categories related to likelihood of sepsis or infection: (1) sepsis (diagnosis including the term "sepsis"); (2) serious infection (infection for which hospitalization and/or surgical or procedural intervention would be deemed necessary, including central nervous system (CNS) infection, intra-abdominal infection, bacteremia, gangrene, neutropenic fever, intrathoracic infection, and orthopedic infection); (3) infection (all other bacterial, fungal, parasitic, or unspecified infections); (4) viral infection (specified viral etiology); (5) organ failure (diagnosis indicating impairment of organ function without reference to etiology); (6) non-infectious (diagnosis specifying a condition not typically associated with infection); and (7) possible infection (diagnosis for which it was not possible to determine if there was an infectious etiology, typically symptom-related). The "infection" category was further classified into seven subcategories of pneumonia; genitourinary (GU) infection; gastrointestinal (GI) infection; skin and soft tissue infection (SSTI); ear, nose and throat (ENT) infection; febrile syndrome; and other or unspecified infection. Table A1 provides examples of diagnoses in each category.

Table B.1: Common and Representative ED Diagnosis by Assigned Diagnosis Category

| Category | Common and representative primary ED diagnoses |
|---|---|
| Infection | |
| Pneumonia | Pneumonia due to infectious organism, unspecified laterality; pneumonia; multifocal pneumonia; pneumonia of right lower lobe due to infectious organism; bacterial pneumonia |
| GU infection | Pyelonephritis; **UTI (urinary tract infection), site unspecified**; **cystitis** |
| GI infection | Colitis; gastroenteritis; cholangitis; stercoral colitis; enterocolitis |

| | |
|---|---|
| SSTI | Cellulitis; cellulitis of left lower extremity; cellulitis of right lower extremity; cellulitis and abscess of leg; cellulitis of lower extremity, unspecified laterality |
| Fever | **Fever, unspecified fever cause**; fever; fever of unknown origin; fever and chills; fever, unknown origin |
| ENT infection | Strep pharyngitis; peritonsillar abscess; strep throat; acute pharyngitis, unspecified etiology; dental infection |
| Other/unspecified Infection | Malaria; wound infection; infection; post-op infection; MRSA infection |
| Serious infection | Neutropenic fever; acute cholecystitis; bacteremia; cholecystitis; diverticulitis of colon; bowel perforation; osteomyelitis; liver abscess; acute bacterial endocarditis; gangrene; meningitis |
| Non-infectious | Cardiac arrest; seizure; **chest pain**; abdominal pain, unspecified abdominal location; syncope and collapse; hypernatremia; obstructive chronic bronchitis with exacerbation; SBO (small bowel obstruction); pleural effusion; **hip pain; constipation; inflammatory disease of uterus** |
| Possible infection | Hypotension, unspecified hypotension type; **shortness of breath**; hypotension; failure to thrive in adult; respiratory distress; weakness; diarrhea, unspecified type; **myalgia** |
| Organ Failure | Altered mental status, unspecified altered mental status type; acute respiratory failure with hypoxia; hypoxia; **AKI (acute kidney injury)**; acute hypoxic respiratory failure; acute respiratory failure with hypoxia and hypercapnia; shock |
| Sepsis | Sepsis, due to unspecified organism, unspecified whether acute organ dysfunction present; sepsis; septic shock; severe sepsis; sepsis without acute organ dysfunction, due to unspecified organism |
| Viral infection | COVID-19; influenza A; **influenza**; acute viral syndrome; respiratory syncytial virus (RSV); upper respiratory tract infection, unspecified type |

Table A2 shows examples of common and representative emergency department diagnoses within categories modeled on the schema of Linden and Melhammar. Categories were assigned by the principal investigator using data obtained in an electronic health record query on 62,068 ED encounters with a sepsis BPA. Bolded diagnoses are those that occurred at the index ED visit among 14 participants who had a 7-day return ED visit with admission for sepsis.

### Appendix C: Blood Culture Results among Participants

Blood cultures were sent during 15,483 participant encounters with hospital admission and 3,026 encounters with ED discharge. Blood cultures were positive in 13.7% of admitted participants, compared with 2.4% of discharged participants. Overall, 14.9% of isolates were likely contaminants or equivocal pathogens—14.8% for admitted participants and 21.9% for discharged participants. The most common likely pathogen species were Escherichia coli (26.7%), Staphylococcus (17.3%), Streptococcus (14.2%), Klebsiella (13.7%), Pseudomonas (4.4%), Enterococcus (3.5%), and Candida (3.3%). Among participants with a return admission for sepsis or infection, blood cultures from the index ED visit were positive in 21 participants (13.4%), with 2 (8.3%) growing likely contaminants.
